# Appropriateness of ChatGPT in answering heart failure related questions

**DOI:** 10.1101/2023.07.07.23292385

**Authors:** Ryan C. King, Jamil S. Samaan, Yee Hui Yeo, Behram Mody, Dawn M. Lombardo, Roxana Ghashghaei

## Abstract

**Introduction:** Heart failure requires complex management with increased patient knowledge shown to improve outcomes. This study assessed the knowledge of Chat Generative Pre-Trained Transformer (ChatGPT) and its appropriateness as a supplemental resource of information for patients with heart failure.

**Materials and Methods:** A total of 107 frequently asked heart failure-related questions were included. Responses were generated using GPT-3.5 and GPT-4. The accuracy and reproducibility of responses were graded by two reviewers board-certified in cardiology, with differences resolved by a third reviewer board-certified in cardiology and advanced heart failure. Accuracy was graded using a four-point scale: 1) Comprehensive 2) Correct but inadequate 3) Some correct and some incorrect 4) Completely incorrect.

**Results:** GPT-4 displayed a greater proportion of comprehensive knowledge for the categories of “basic knowledge” and “management” (89.8%, 82.9%). GPT-3.5 performed best in the “management” and “other” category (prognosis, procedures, and support) (78.1%, 94.1%). There were two total responses (1.9%) graded as “some correct and incorrect” for GPT-3.5, while no “completely incorrect” responses were produced. The models also provided highly reproducible responses, with GPT-3.5 scoring above 94% in every category and GPT-4 with 100% for all answers.

**Conclusions:** GPT-3.5 and GPT-4 answered the majority of heart failure-related questions accurately and reliably. ChatGPT may lead to better outcomes in patients with heart failure by providing accessible and easy to understand health education. However, in its current state the model necessitates further rigorous testing and validation to ensure patient safety and equity across all patient demographics.

## Introduction

Heart failure is a chronic condition with a healthcare burden forecasted to grow to approximately $70 billion annually in the US by 2030. This cost is predominantly constituted by hospitalizations (70%) which represent 1-2% of total US hospital admissions [1]. Enhanced patient understanding of heart failure management has been shown to lead to fewer and shorter duration of hospital admissions [2]. In search of answers, patients often utilize online resources for information regarding their health resulting in approximately one billion daily google searches related to healthcare [3].

The large language model (LLM), Chat Generative Pre-Trained Transformer (ChatGPT), is an artificial intelligence (AI) model that was trained on a large dataset comprising a vast spectrum of topics including medicine. It provides text-based responses in a conversational manner to questions prompted by users [4]. Beyond its rapid growth in popularity, the model continues to improve in performance with the latest version, GPT-4, released in March of 2023, which has been shown to significantly outperform its predecessor, GPT-3.5 across multiple tasks and domains [5]. As ChatGPT’s capabilities grow, we hypothesize patients will be more inclined to seek information from this technology regarding complex conditions such as heart failure.

The model’s utility in medicine is actively under investigation with prior studies having examined ChatGPT’s ability to answer questions related to heart disease prevention, bariatric surgery, and cirrhosis yielding promising results [6,7,8]. In the deployment of tools such as ChatGPT within the medical field, a comprehensive examination of both strengths and limitations is essential. Furthermore, informing patients of ChatGPT’s potential benefits and shortcomings can facilitate shared decision-making and align expectations, thus enhancing the overall healthcare experience. To better understand the potential role of ChatGPT as an adjunct source of information for patients with heart failure, we examined the accuracy and reproducibility of ChatGPT’s responses to heart failure related questions. Furthermore, we assessed the difference in performance between GPT-3.5 and GPT-4.

## Materials and Methods

We curated a list of 107 frequently asked questions related to heart failure from medical societies, renowned medical institutions, and Facebook support groups. Questions were entered twice into each model (GPT-3.5 and GPT-4) using the “new chat” function, producing two responses per question per model. Responses were first graded independently by two board-certified cardiologists actively practicing in a tertiary academic center. Accuracy was graded using the following scale: 1) Comprehensive 2) Correct but inadequate 3) Some correct and some incorrect 4) Completely incorrect. Reproducibility of responses was determined by categorizing responses into two categories based on the grading of each response. Categorization was determined by the presence of incorrect information in the response. The first category included scores of “comprehensive” and “correct but inadequate” while the second category comprised scores of “some correct and some incorrect” and “completely incorrect”. A question with its responses in different categories, thereby difference in the accuracy of the responses, was defined as non-reproducible. Discrepancies in grading between the two reviewers were resolved by a third, senior reviewer board-certified in advanced heart failure and actively practicing in a tertiary academic center.

### Statistical analysis

Descriptive analysis includes proportions shown as counts and percentages. Microsoft Excel (version 16.68) was used for all statistical analysis.

## Results

The majority of responses from both models were graded as either “comprehensive” or “correct but inadequate” (**Table 1**). GPT-4 provided 107/107 (100%) responses with correct information with 89/107 (83.2%) of responses graded as “comprehensive” while GPT-3.5 provided 105/107 (98.1%) responses with correct information and 84/107 (78.5%) of responses graded as “comprehensive”. GPT-4 performed best in the categories of “basic knowledge” and “management” producing comprehensive responses 89.8% and 82.9% of the time, respectively. Its predecessor, GPT-3.5, displayed its most comprehensive responses for questions related to “management” (78.1%) and the category of “other” (94.1%). The models provided a thorough response regarding the benefits of cardiac rehabilitation mentioning factors such as reduced risk of future complications leading to additional hospitalizations and higher healthcare costs, increased medication adherence, and patient education along with support. When examining presence of incorrect information among responses, GPT-4 provided no inaccurate information while GPT-3.5 did. GPT-3.5 provided two responses (1.9%) which were graded as “some correct and some incorrect”, while neither model produced “completely incorrect” responses. GPT-3.5 incorrectly stated that having heart failure inherently increases the risk of experiencing a myocardial infarction without including that some types such as infiltrative cardiomyopathies may not carry as high of a risk. When examining reproducibility, the models provided reproducible responses for the majority of questions, with GPT-3.5 scoring above 94% in every category and GPT-4 with 100% for all responses (**Table 2**). A distinction between models was noted regarding the depth of detail in responses for certain questions. For example, GPT-3.5 answered in general terms regarding the cardiac benefits of SGLT2 inhibitors, while GPT-4 provided a more detailed yet succinct response regarding their effects on diuresis and blood pressure.

**Table 1.** Summary of grading of responses produced by GPT-3.5 and GPT-4.0 to heart failure related questions organized by topic.

| **Grading Scale** | **GPT-3.5** | **GPT-4** |
| --- | --- | --- |
| *Overall (N=107)* | Number of responses (%) |  |
| 1. Comprehensive | 84 (78.5) | 89 (83.2) |
| 2. Correct but incomplete | 21 (19.6) | 18 (16.8) |
| 3. Some correct and some incorrect | 2 (1.9) | 0 (0.0) |
| 4. Completely incorrect | 0 (0.0) | 0 (0.0) |
| *Basic Knowledge (N=49)* |  |  |
| 1. Comprehensive | 36 (73.5) | 44 (89.8) |
| 2. Correct but incomplete | 13 (26.5) | 5 (10.2) |
| 3. Some correct and some incorrect | 0 (0.0) | 0 (0.0) |
| 4. Completely incorrect | 0 (0.0) | 0 (0.0) |
| *Management (N=41)* |  |  |
| 1. Comprehensive | 32 (78.1) | 34 (82.9) |
| 2. Correct but incomplete | 8 (19.5) | 7 (17.1) |
| 3. Some correct and some incorrect | 1 (2.4) | 0 (0.0) |
| 4. Completely incorrect | 0 (0.0) | 0 (0.0) |
| *Other-prognosis, procedures, support (N= 17)* |  |  |
| 1. Comprehensive | 16 (94.1) | 11 (64.7) |
| 2. Correct but incomplete | 0 (0.0) | 6 (35.3) |
| 3. Some correct and some incorrect | 1 (5.9) | 0 (0.0) |
| 4. Completely incorrect | 0 (0.0) | 0 (0.0) |

**Table 2.** Reproducibility of responses by GPT-3.5 and GPT-4.0 to heart failure related questions categorized by question subgroup.

| Question subgroup | GPT-3.5 | GPT-4 |
| --- | --- | --- |
|  | Number of responses (%) |  |
| **Overall (N=107)** | 105 (98.1) | 107 (100.0) |
| **Basic Knowledge (N=49)** | 49 (100.0) | 49 (100.0) |
| **Management (N=41)** | 40 (97.6) | 41 (100.0) |
| **Other (N= 17)** | 16 (94.1) | 17 (100.0) |

## Discussion

We examined the accuracy and reproducibility of responses by GPT-3.5 and GPT-4 to questions related to heart failure. GPT-4 provided comprehensive responses to 83.2% of questions overall while producing no questions containing inaccurate information. On the other hand, GPT-3.5 produced 78.5% comprehensive responses and two (1.9%) containing incorrect information. The models also provided reproducible responses to the majority of questions overall (GPT-4 100%, GPT-3.5 98.1%). The results of our study show ChatGPT’s ability to reliably provide comprehensive responses to patients’ questions. Our findings highlight the potential future of LLMs as an accurate and reliable resource for patients with heart failure to use in addition to care provided by their healthcare provider.

ChatGPT may lead to better outcomes through patient empowerment with knowledge as seen in prior studies investigating the effect of health education on heart failure management [2]. We anticipate patients will seek answers related to their health from ChatGPT due to its simple user interface and easy-to-understand human-like responses provided in a conversational format. GPT-4’s high rate of accuracy can likely be attributed to its training which focused on better understanding of users’ intent and processing of more complex scenarios [5]. The performance of ChatGPT in this study shows that this emerging technology may be a useful tool for both patients and providers in the future but does require further rigorous testing and validation. We recommend that researchers and clinicians continue investigating the capabilities and limitations of ChatGPT and other AI platforms to maximize its impact on improving patient outcomes.

Future research analyzing ChatGPT may benefit from considering challenges and limitations involved in this study and within the model itself. For example, the reviewers in this study were unable to be blinded to the identity of each ChatGPT version for a specific response due to the unexpected release of GPT-4 during the study period. Although a grading system involving a panel of multiple reviewers was used, bias may still exist through subjective review. ChatGPT performed well in this study with few incorrect responses, but there are additional intrinsic concerns that require improvement before it is suitable to significantly contribute to medical management. On occasion, the model may respond with inaccurate information that is organized in a believable manner that may be deceiving due to its human-like text. Nonsensical responses may potentially be produced as well [4]. Furthermore, the accuracy of the model depends on an undisclosed dataset only including information prior to September 2021 [4]. Bias may also inherently be introduced as a programmer guides the conversation with ChatGPT during the reinforcement learning from human feedback (RLHF) training phase. The consistency of recommendations may also vary from region to region. Given these challenges, we must act before the technology becomes too advanced to regulate effectively.

The rapid evolution of AI, especially LLMs like ChatGPT, calls for in-depth ethical and regulatory discussions for healthcare applications. These models generate medical content based on prior scientific research, but the exact sources of input remain unknown raising questions of credibility and patient safety [9]. There are also concerns of patient privacy. Open AI recently added an option to disable the chat history allowing a user to prevent a conversation from being used in the model’s training [10]. However, there is no consent or explanation describing how a user’s personal health related queries will be used in this process or beyond. Comprehensive oversight is needed to address these ethical concerns while also promoting transparency and prioritizing patient safety. The Food and Drug Administration (FDA) has proposed regulating these AI technologies similar to medical devices through continuous monitoring and recurrent validation [11]. Such regulation would ensure that these dynamic tools are appropriately validated for their intended use and are continuously reevaluated for safety and performance.

Upon rigorous evaluation and approval, ChatGPT offers promising healthcare applications that may streamline efficiency and improve clinical practice. The online medical platform Doximity recently released a beta website, designed with physician input, that uses ChatGPT for administrative tasks such as faxing preauthorization documents and insurance appeals while maintaining HIPAA compliance [12]. For future applications, it is crucial that physicians remain actively involved in the design process to maintain trust and safety. ChatGPT has also shown promise in writing medical notes in an ICU setting when provided with various clinical information. It performed well when organizing objective data, but encountered challenges in making connections between complex concepts often present in multisystem disease processes [13]. This underscores credibility and reliability concerns that regulatory bodies must address if ChatGPT is to guide complex medical decisions in the future.

Although the technology holds exciting potential, the healthcare community must be vigilant to prevent existing disparities from perpetuating and worsening. As AI systems like ChatGPT become more prevalent in healthcare, ensuring their equitable development and deployment is vital. The FDA’s Drug Trials Snapshots program highlights the underrepresentation of minorities in clinical trials from 2015-2019 compared to the US census, while white participants were notably overrepresented [14]. If ChatGPT’s training data leans towards specific demographic groups, it risks perpetuating bias, offering less relevant advice for marginalized groups. If these biases are addressed and the model is validated, then this technology holds incredible promise in bridging disparities present in lower socioeconomic healthcare settings such as resource-constrained free clinics as proposed by Ong, et al [15]. ChatGPT may help enhance patient care by providing transportation and housing resources, creating appropriate patient education materials, and overcoming language challenges. Its multilingual capability, which has shown remarkable accuracy in several non-English languages, can facilitate real-time multilanguage discussions and assessments of a patient’s relevant medical history to be further reviewed by a healthcare professional [15-17]. While ChatGPT’s integration into healthcare presents challenges due to potential biases, its proper validation and application could revolutionize patient care offering a promising avenue for a more inclusive and effective healthcare future.

## Conclusion

GPT-3.5 and GPT-4 provided accurate and reliable responses to the majority of heart failure-related questions in this study, with notable superior performance from GPT-4 by providing no incorrect information in its responses. The performance of GPT-4 demonstrates the impressive ongoing improvement in LLM technology and highlights the future potential of LLMs as adjunct sources of information for patients with heart failure. However, in its current state the technology still requires rigorous testing and validation by regulatory medical bodies to ensure both patient safety and equitable access to healthcare resources. We recommend future investigation into the capabilities and limitations of ChatGPT and other AI platforms to identify their impact on patient outcomes.

## Data Availability

All data produced in the present study are available upon reasonable request to the authors

## Notes

### Competing Interest Statement

The authors have declared no competing interest.

### Funding Statement

This study did not receive any funding

### Summary of Updates

The revised version of this manuscript addresses the limitations and ethical concerns of ChatGPT more comprehensively than prior versions.

